# Placental telomere length shortening is not associated with preeclampsia but the gestational age

**DOI:** 10.1101/2022.02.28.22271630

**Authors:** Xiaotong Yang, Paula A Benny, Elorri Cervera-Marzal, Biyu Wu, Cameron Lassiter, Joshua Astern, Lana X. Garmire

**Affiliations:** Department of Computational Medicine and Bioinformatics, University of Michigan, Ann Arbor, Michigan, United States of America; Epidemiology Program, University of Hawaii Cancer Center, Honolulu, Hawaii, United States of America; Department of Nutritional Science, University of Hawaii, Honolulu, Hawaii, United States of America; University of Hawai’i Biorepository, Burns School of Medicine, University of Hawaii at Manoa, Honolulu, United States of America

**Keywords:** Preeclampsia, placenta, telomere length, gestational age, ethnicity, sex, blood type

## Abstract

**Background:** Preeclampsia is a severe pregnancy complication that affects about 2-8% of pregnant women globally and is one of the leading causes of maternal and fetal mortality and morbidity. Telomere length change has been associated with aging, environmental stress, and various diseases. Studying the telomere length change in preeclampsia complicated placentas may shed light on the pathophysiology of preeclampsia.

**Objectives:** In this study, we aimed to explore if associations exist between placental telomere length and preeclampsia, gestational age, and other demographic, clinical and physiological factors related to pregnancy.

**Study Design:** We collected placentas samples from 120 severe preeclamptic deliveries and 129 healthy full-term deliveries in Hawaii between year 2006 and 2013 and measured the average absolute placental telomeres with quantitative polymerase chain reaction (qPCR). We retrieved their pre-recorded demographic, clinical and physiological data and conducted multiple linear regressions to associate placental telomeres with preeclampsia and other variables.

**Results:** Placental telomere length of severe preeclampsia cases showed no significant difference (P=0.20) from healthy controls after controlling for gestational age difference. Instead, we identified that placental telomere length consistently decreases as gestation progresses (P=6.10e-05). Male babies have shorter placental telomere than female babies in the early trimester (p = 0.024), but the placentas of male babies have a reduced rate of telomere shortening along pregnancies, compared to those of female babies (p = 0.029). Latino mothers show longer placenta telomeres than other ethnicities (P=0.012), whereas mothers of blood type O have shorter placenta telomeres than those of other blood types (P=0.039).

**Conclusions:** Using the largest multiethnic cohort to date, we showed the lack of association between preeclampsia and placental telomere length. Rather, gestational age is the most dominant variable associated with placental telomere shortening.

**Condensation:** Placental telomere length shows no significant association with preeclampsia but significantly shortens with the progressing gestational age at different rates for male and female babies.

**AJOG At A Glance:** *Why was this study conducted?:*

- Understanding the TL change in healthy and preeclampsia complicated placenta may help us better understand the physio-pathological impact of preeclampsia.

*What are the key findings?:*

- We found no significant association between severe preeclampsia and placental TL in the largest multi-ethnic cohort to date.
- Placental TL shortens consistently largely due to gestational age increase.
- Placentas of male babies have shorter TLs at the early third trimester but slower TL shortening compared to those of female babies.
- Latino mothers have longer placental TL compared to non-Latino mothers.
- Mothers of blood type O have shorter placenta TL than others.

*What does this study add to what is already known?:*

- We showed that placental telomere length shortening is not associated with preeclampsia but the gestational age in a large multi-ethnical cohort.

## INTRODUCTION

Preeclampsia (PE) is a pregnancy syndrome in women after 20 weeks of pregnancy.^1^ The two major clinical symptoms are gestational hypertension (systolic blood pressure >= 140 mm hg or diastolic blood pressure >= 90) and proteins in urine. Other clinical symptoms include renal failure, cardiac dysfunction or arrest, stroke, and even maternal and fetal fatalities.^1^ The average incidence rate of preeclampsia is about 2-8% globally and 3.1% in the United States.^2,3^

Preeclampsia is one of the leading causes of pregnancy-related mortality and is associated with many long-term maternal and fetal morbidities. Major risk factors of preeclampsia include prior preeclampsia, chronic hypertension, multiple gestation, higher body mass index (BMI), pregestational diabetes, and antiphospholipid syndrome.^4^ Preeclampsia is an intricate and heterogeneous disorder containing multiple subtypes: based on the severity of the symptoms, preeclampsia can be classified as mild, severe or superimposed^1^; based on the occurrence time, preeclampsia that appears before week 34 of gestation is considered early-onset preeclampsia and after week 34 is called late-onset preeclampsia^5^. Epigenetic, transcriptomic, proteomic, and metabolic approaches were adapted in preeclampsia studies recently, however, the molecular mechanisms leading to these symptoms are still not fully understood.^6,7^

Telomeres consist of repeated DNA sequences and protective protein complexes. They function as the protective structures at chromosome ends.^7^ Telomeres shorten continuously in each cell division and the process may be accelerated by chorionic diseases and environmental stress.^9^ The telomere length (TL) contains important information about cellular state, and it has been considered a biomarker for aging^10^. Placentas play a central role in PE, and it is believed that the onset of PE originated from defective placentation, and the release of antiangiogenic factors from the placenta into the maternal circulation. Previous epigenetic studies have suggested accelerated aging in placentas from early-onset PE^11^. Therefore, it is of great interest to investigate if such presumably “pre-aging” stress exerted in placenta epigenetics is also manifested in TL shortening. However, various other factors may significantly confound placenta TL interpretation, such as gestational age, ethnicities, the geolocation of the tissue samples taken from the placenta. Failure to address these important confounders may have contributed to the previous controversy regarding the association between PE and placental TL^12,13,14^. To resolve this issue, we conducted the largest multiethnic cohort (MEC) study so far, containing placentas of 120 severe PE and 129 matched full-term deliveries from the Hawaii Biorepository (HiBR). The placenta tissues were obtained from deliveries in Kapiolani Women and Children’s Hospital in Honolulu, Hawaii between January 2006 and June 2013. We measured the total placental TL by qPCR technology, and then conducted linear regression analysis between placental TL and physiological and clinical factors. Our objectives are: (1) studying if there is indeed an association between severe PE and placental TL shortening; (2) revealing other potential maternal or fetal factors associated with placental TL.

## MATERIAL AND METHODS

### Biospecimens

Biospecimens were obtained from the Hawaiian Biorepository (IRB #CHS23976). Placenta samples were collected from the intermediate portion of the placenta after maternal and fetal membranes were trimmed away. Samples with neonatal malformation were removed from the analysis. As a result, samples from 249 women (120 with severe PE and 129 full-term healthy controls), who delivered their babies in Hawaii between January 2006 and June 2013 were collected and analyzed in this study. Severe PE was defined as sustained pregnancy-induced hypertension with urine protein and/or organ dysfunction according to the diagnosis of OBGYNs in Kapiolani Medical Center. For each case, a matched control sample was identified using mother age, prepregnant body mass index (BMI) and ethnicity group as the matching criteria.

### Average absolute TL measurement

The placenta tissue samples were stored at −80°C. The genomic DNA extracted from placenta tissue was performed using AllPrep DNA/RNA/Protein Mini Kit (Qiagen, USA) according to the manufacturer’s instructions. Briefly, 30mg of frozen placenta tissue samples were weighted in a 50 ml Falcon tube (VWR, USA) and mixed with 600 μl buffer RLT (Qiagen, USA) supplemented with 1% β-mercap-toethanol (Sigma). The tissue was then homogenized with Tissue Ruptor II (Qiagen, USA) for 30 seconds at medium speed. The lysate was transferred to a 2 ml microcentrifuge tube and centrifuged for 3 minutes at 13,000g at 4 °C. Then the supernatant was pipetted into an AllPrep DNA spin column (Qiagen) and processed following Kit’s instruction. The DNA concentration and purity were evaluated by NanoDrop2000 spectrophotometer (Thermo Scientific, Waltham, USA) and the DNA integrity was investigated by electrophoresis in 1% agarose gel with ethidium bromide (Sigma).

The average TL of placenta tissue samples was measured using the Absolute Human TL Quantification qPCR assay Kit (AHTLQ, ScienCell Research Laboratories, USA). Each sample was tested in duplicate with StepOne™ Real Time System (Applied Biosystems, USA) using 96-well reaction plates. The melting curve was observed to evaluate the specificity and efficiency of the qPCR reaction. The Comparative ΔΔCq (Quantification Cycle Value) method was applied to calculate the relative average TL of target tissue samples^15^. A 100 bp region single copy reference (SCR) primer set on chromosome 17 was used as the reference. The absolute TL is calculated as the relative TL multiplied by the SCR TL.

### Demographical, clinical and physiological data

A total of 20 demographical, clinical and physiological variables obtained from the University of Hawaii HiBR biobank were used in this study: mother’s age, height, pre-pregnancy BMI, weight gain during pregnancy, blood type (A vs non-A, B vs non-B and O vs non-O), ethnicity (Asian vs non-Asian, Caucasian vs non-Caucasian, Latin vs non-Latin and Pacific Islander vs non-Pacific Islander), history of asthma, history of anemia, chronic hypertension, macrosomia, membrane ruptured, vaginal bleeding, babies’ sex, gestational age in weeks, and sample group. The missing values were imputed with the predictive mean match (PMM) algorithm from R package “*mice”*^*16*^.

### Statistical analysis

Two-sided Welch t-tests and Chi-squared tests were used to compare numeric and categorical characteristics between case and control groups. Logistic regression was performed to measure the correlations between severe PE and other demographical and clinical variables. The proportional variance explained (PVE) values of all variables were calculated using eta-values from the 1-way ANOVA result. The PVE value describes the proportion of total variance in data that can be explained by a certain variable, demonstrating its importance^17^. The absolute telomere length was ln-transformed to reduce skewness and ensure normality. TL refers to ln-transformed placental TL unless stated otherwise.

A multiple linear regression model was constructed by regressing TL of 249 samples on all 20 variables and the most significant interaction between gestational age and infant sex. We refer to this model as the full model. To select the most relevant variables and improve model fitness, we applied the stepwise selection algorithm on the full model to search for the optimal model with the lowest Akaike Information Criterion (AIC), by adding or removing one variable at a time. This stepwise selected model is referred to as the optimal model, including 7 variables: gestational age, history of Asthma, Baby’s sex, Blood type O, Latino ethnicity, and the interaction between baby’s sex and gestational age.

All analyses were conducted with different packages in R software version 4.0.2.^18^ “*tidyr*”, and “*dplyr*” were used in data cleaning; “*mice*” was used to impute missing values; “*stats*” and “*MASS*” was used in statistical analysis; “*ggplot2*”, “*gridExtra*”, “*ggsignif*” and “*corrplot*” were used in visualization^16,19–25^. All R script used in this project is in repository.

## RESULTS

### Overview of the study cohort

We present the phenotypic characteristics of 120 pregnancies with severe PE and 129 healthy controls in Table 1. There is no significant difference in age, height, BMI, blood type, ethnicity and weight gain between the severe PE cases and controls, confirming the success of matching criteria in the study design. Significantly more mothers from the severe PE group have other pre-existing conditions including the history of anemia (p = 0.024) and chorionic hypertension (p = 5.58e-05). More mothers from the control group have experienced vaginal bleeding (p=9.70e-3) and the rupture of membrane (p=7.30e-3). Babies from the control group are more likely to have macrosomia (p = 0.023). The PE group has a much smaller gestational age (p < 2.83e-29): the average and standard deviation of the gestational age in severe PE cases is 35.35 weeks +/− 2.90 weeks, compared to controls 39.33 weeks +/− 0.85. Further relationships between clinical variables can be found in the Pearson’s correlation heatmap of all phenotypic variables (Supplementary Figure 1).

**Table 1:** Differences in demographic and clinical variables between severe PE cases and controls.

| *Variables* | *Preeclampsia (n = 120)*<br>*Mean(sd)* | *Control (n = 129)*<br>*Mean(sd)* | *p-value\**<br>*(Two group)* |
| --- | --- | --- | --- |
| *Gestational Age (week)* | 35.36(2.90) | 39.33 (0.85) | 1.76e-29<br>*** |
| *Mother's Age* | 29.81(6.54) | 29.53(6.53) | NS |
| *Weight Gain (lbs)* | 34.17(19.62) | 31.80(16.21) | NS |
| *Mother's Height* | 5.21(0.47) | 5.30(0.38) | NS |
| *Pre-pregnancy BMI* | 29.10(7.53) | 28.01(6.19) | NS |
| *Blood Type (n)* |  |  | NS |
| A | 34 | 43 | - |
| AB | 11 | 9 | - |
| *B* | 29 | 26 | - |
| *O* | 46 | 51 | - |
| ***Ethnicity (n)*** |  |  | NS |
| *African American* | 2 | 2 | - |
| *Asian* | 62 | 72 | - |
| *Caucasian* | 15 | 17 | - |
| *Pacific Islander* | 32 | 27 | - |
| *Latin* | 10 | 11 | - |
| ***Ln Telomere Length*** | 5.58 (0.42) | 5.41(0.38) | 1.18e-3 *** |
| ***Vaginal Bleeding (n)*** | 10 | 32 | 9.70e-3*** |
| ***Macrosomia(n)*** | 2 | 11 | 0.032* |
| ***Chronic Hypertension (n)*** | 35 | 11 | 5.58e-05*** |
| ***History of Asthma (n)*** | 21 | 19 | NS |
| ***History of Anemia (n)*** | 28 | 15 | 0.023* |
| ***Baby sex (n)*** |  |  | NS |
| *Male* | 69 | 61 | - |
| *Female* | 51 | 68 | - |
| ***Membrane ruptured(n)*** | 23 | 51 | 7.38e-3*** |

\*Categorical variables are compared with Chi-square test. Numeric variables are compared with t-test.
\* Significance level: \* <0.05, \*\* <0.01, \*\*\*<0.001

The distribution of absolute TL is highly skewed with a long right tail (Supplement Figure 2A). To approximate the Gaussian distribution of linear regression, we ln-transformed TL, like others^12^. All the TL in the remaining report refers to ln-transformed TL unless noted otherwise. Ln-transformed TL has a bell-shaped distribution with values ranging from 4.55 to 6.73 (Figure 1A). The average TL in PE cases is 5.59, significantly higher than the average value of TL in controls 5.41 (p = 7.18e-4) (Table 1).

**Figure 1:**
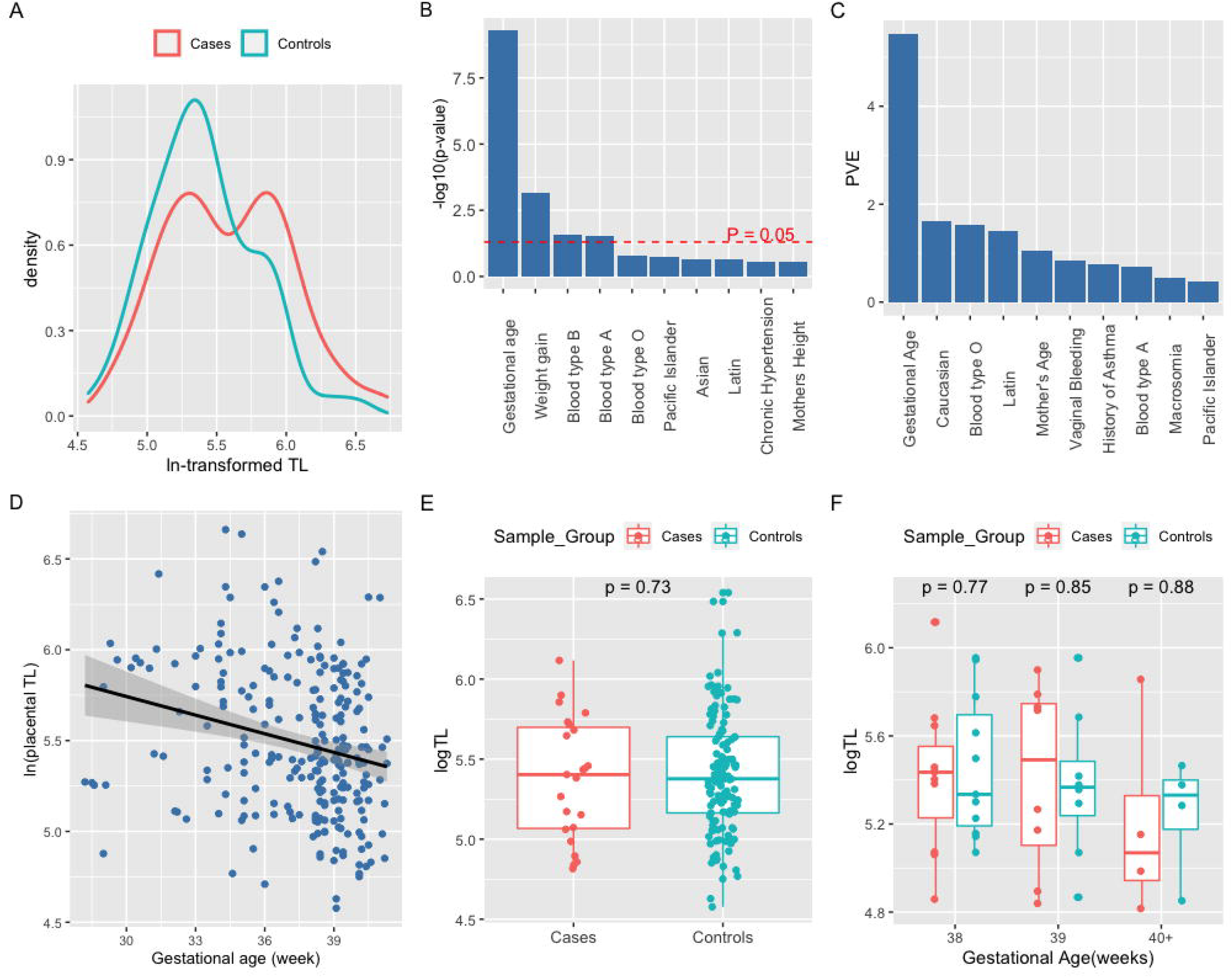
The lack of association between severe PE and TL after stratifying by gestational age. **A**: Distribution of ln-transformed relative placental TL in the third trimester. Blue color: severe PE samples; Red color: control samples. **B**: Top 10 most relevant phenotypic variables to fit severe PE by logistic regression. **C**: Top 10 variables with the highest proportion of variance explained (PVE), per ANOVA analysis. **D**: Scatter plot of TL by gestational age, the solid line represents linear regression of TL on gestational age. **E**: Boxplots of placenta TL of term PE samples (n=29) and term controls (n=133). **F**: Boxplots of placenta TL of term PE samples (n=29) and the subset of matched term controls (n=29) stratified by gestational age (38, 39, 40+ weeks).

**Figure 2:**
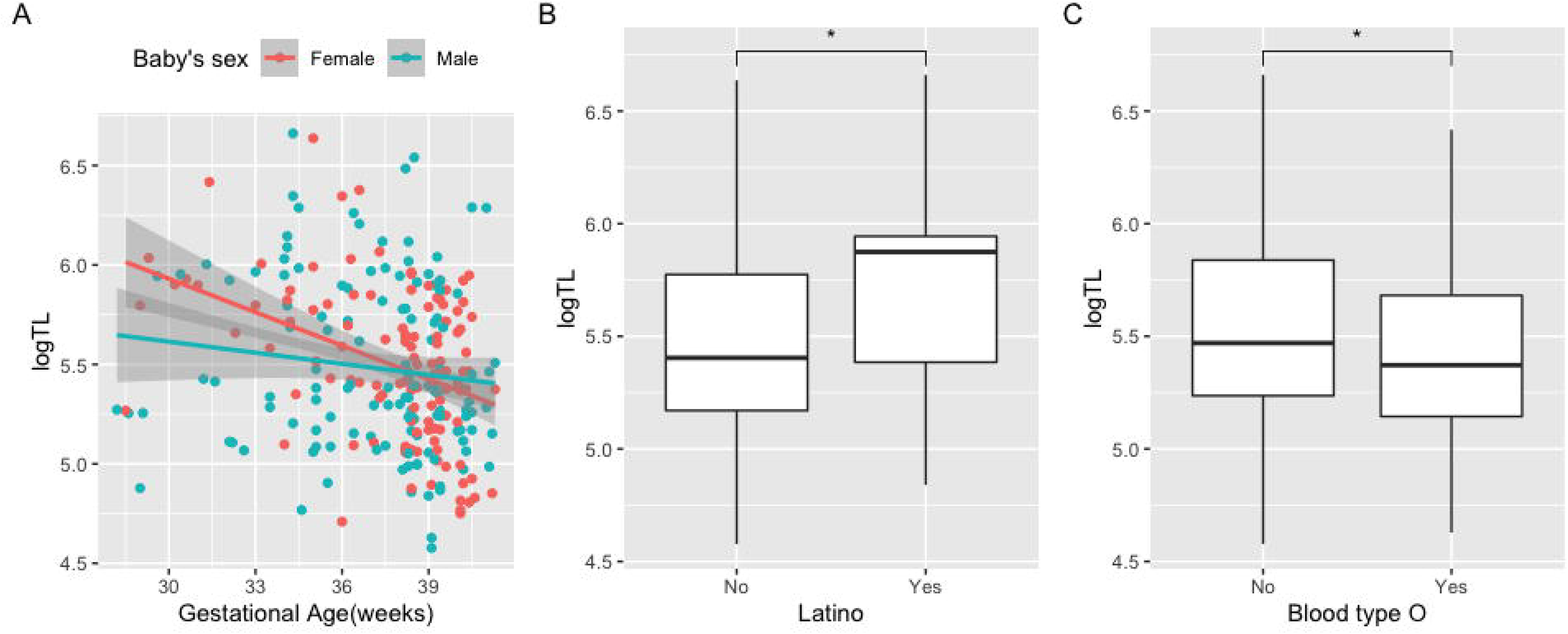
The associations between placental TL and other demographic and clinical variables. **A**: The interactive effect of baby sex and gestational age on placental TL; red color: female, blue color: male. Red and blue lines: linear regression of TL over gestational age for female (red) and male babies (blue). **B**: The boxplot of placental TLs in Latino mothers vs non-Latino mothers. **C**: The boxplot of placental TL in mothers of blood type O vs mothers of non-O blood type.

### Gestational age, instead of severe PE, shows significant association with TL

Although placental TL is longer in the PE group from the exploratory analysis (Table 1), such difference may be due to other confounding factors, such as gestational age. Indeed, the logistic regression model using 19 other clinical variables to fit PE shows that gestational age has the strongest relationship with PE (log_10_(p-value) = 9.29, p = 5.07e-10), among other 3 variables that have significant correlations with PE: weight gain (log_10_(p-value) = 3.15, p = 7.1e-3), blood type B (log_10_(p-value) = 1.55, p = 0.028) and blood type A (log_10_(p-value) = 1.51, p = 0.031) (Figure 1B). Moreover, when examining the contribution of all 20 phenotypic variables to TL variation, gestational age shows the highest and most dominant proportion variance of 5.49% explained according to the ANOVA test, much higher than that of 1.65% from the 2nd ranked Caucasian ethnicity, while severe PE is not ranked among the top 10 variables (Figure 1C). Thus, the exploratory analyses coherently show the important relationship between gestational age and TL change in the third trimester.

To quantitatively delineate the relationship between TL and phenotypic factors including severe PE, we then constructed a full linear regression model by regressing placental TL on all 20 demographic and clinical variables and significant pairwise interactions among them, using both PE and control samples (n=249). This full model shows no significant difference (P = 0.20) in TL between PE patients and controls (Supplementary Table 1). We next determined the optimal model (Table 2), by applying the stepwise selection algorithm with the lowest Akaike Information Criterion (AIC) on the full model. Again, PE was excluded from the optimal model due to its lack of contribution to model fitness. Instead, placental TL shows a negative association with gestational age (beta = −0.054, p = 6.10e-5) according to the optimal model (Figure 1D, Table 2). The consistent decrease in TL is correlated with the active cell divisions in the placenta through the last trimester of pregnancy.

**Table 2:**
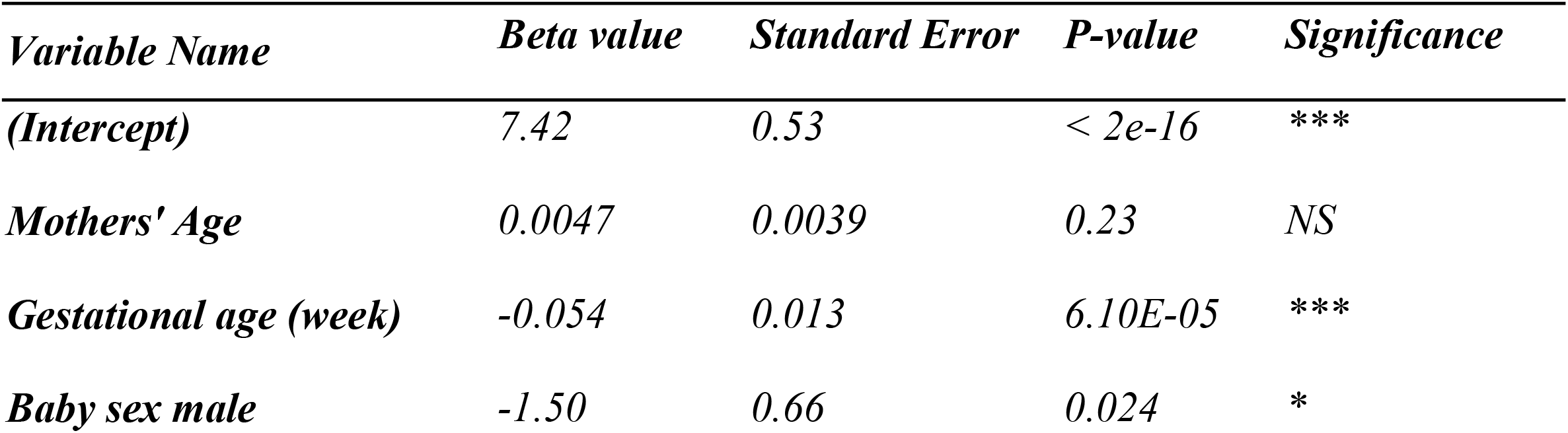
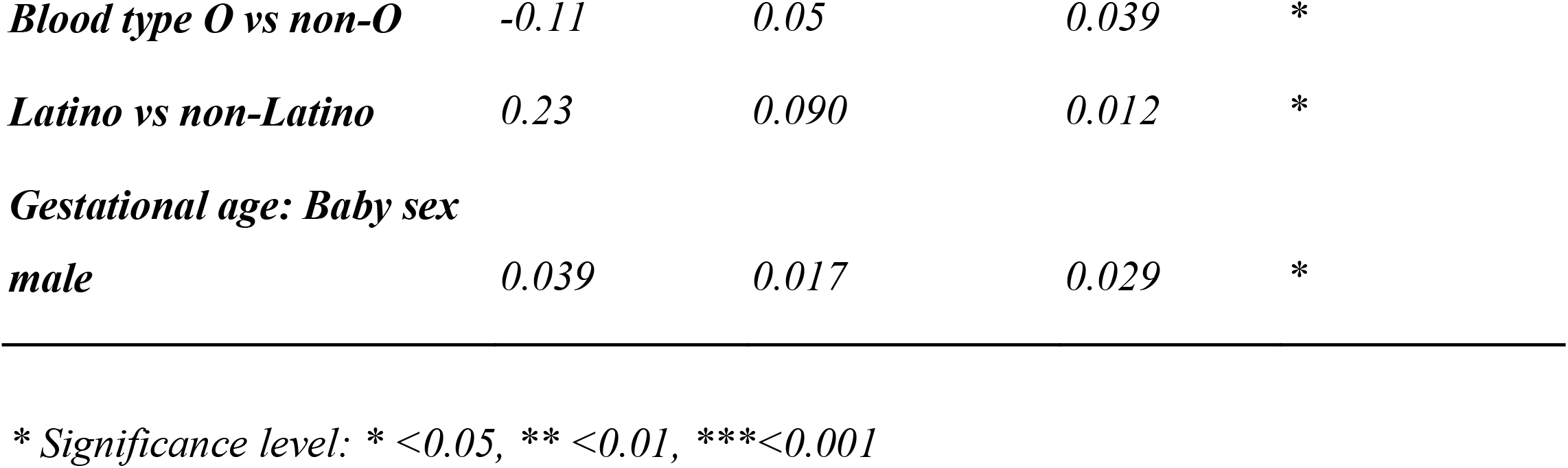
The optimal linear regression model results using 249 samples.

To better control the confounding effect from gestational age on TL, we next focused on the subset of 29 PE patients who reached term gestational ages (>= 38 weeks), same as the 129 control samples. Boxplot shows no significant difference (P =0.73) in TL between full-term PE and controls (Figure 1E). Further, we constructed a linear regression model of TL on all 20 phenotypic variables on this subset (Supplementary Table 2). Again, it shows a lack of association between PE and TL (P =0.65). Additionally, we performed a gestational week stratified (weeks 38, 39 and 40+) comparison between full-term PE samples and their controls, who are matched on gestational age, ethnicity, mother’s age and BMI. Again, boxplots show no significant difference (P = 0.77, P= 0.85, and P= 0.88) of TL between cases and controls in all gestational week strata (Figure 1F). In all, our analyses from many different perspectives all show that the shorter telomeres observed in severe PE patients are mostly associated with the gestational ages rather than PE itself.

### Placental TL and infant sex, maternal ethnicity and maternal blood type

Besides gestational age, the optimal model also revealed significant associations of placental TL to 6 other demographic or clinical variables: male baby (beta = −1.496, P = 0.024), interaction between male baby and gestational age (beta = 0.0386, P=0.029), Latino ethnicity (beta = 0.228, P= 0.012), and blood type O (beta = −0.106, P=0.039) (Table 3).

The association between placental TL and babies’ sex is very interesting. Male babies have shorter placental TL at the beginning of pregnancies, but their TLs decrease at a slower rate compared to those of female babies. At gestational week 30, the log-transformed placental TLs of male babies is estimated 0.32 shorter than those of female babies. The difference keeps getting smaller as gestation progresses, and by week 38.7 male babies’ estimated TLs are the same as those of female babies (Figure 2A). Oppositely, at week 40 the estimated TLs of male babies are even longer than that of female babies. Among mothers of different ethnicities, Latino mothers have longer placental TL than non-Latino mothers after adjusting for age difference (Figure 2B); no significant difference was found among placental TLs from Asian, Caucasian and Pacific Islander mothers. Intriguingly, mothers with blood type O have shorter TL compared to mothers with other blood types (Figure 2C).

## COMMENTS

### Principal Findings

The most major finding of this study is that placental TL shortening is not significantly associated with severe PE; rather, it shortens significantly as gestational age increases. Male babies have shorter placental TL in the early third trimester compared to female babies, but their TLs decrease at a slower rate. Latino mothers have longer placental TLs compared to non-Latino mothers. Mothers of blood type O have shorter TLs than mothers of other blood types.

### Results and Implications

Our data analysis shows the lack of association between severe PE and placental TL, even though the severe PE samples do have longer TL. This apparent longer placenta TL in PE is mostly attributed to earlier gestational ages that accompany the preterm deliveries of babies of PE patients^26^. We have shown here that the dominant contributor for placenta TL shortening is gestational age, rather than the PE condition itself. When the placenta grows with the gestational age, cells divide and telomeres shorten^27^. Support to this conclusion comes from the quantitative multiple regression model, where the other potential confounders are considered. The lack of association between PE and placental TL agrees with several previous findings, where TL was measured by qPCR and telomere restriction fragments (TRF) method.^13,14, 27^ The only study showing a negative association between PE and placental TL had a smaller sample size (14 PE cases, 20 controls).^12^ This study also didn’t report or adjust for other variables, though gestational age (36 +/− 1.41 for the PE, 37.15+/−3.9 for the controls) is less of a concern. It also used a different method Fluorescence in situ hybridization (FISH) for TL measurement. These differences in measurement approaches and statistical rigor may contribute to their different conclusions.

Another interesting finding is the slower TL decrease in placentas of male babies. Many previous studies have explored the relation of infant sex and placental, cord blood, or fetal leukocyte TLs, yet didn’t reach a consensus on the significance of this relation^28–31^. The conflict may originate from the observed interactive effect of infant sex and gestational age on TL. Studies with larger variation in gestational age tend to report longer TL in female infants, which agrees with our finding that male infants have shorter TL until late in the third trimester^28–30^. The study that found no difference in TL between infant sexes had a very small standard deviation in gestational age (sd = 0.15)^31^. The short gestation age window may be responsible for such a conclusion.

Latino mothers were found with longer TL compared to non-Latino mothers after age adjustment, while no significant differences in TL were found among other ethnic groups. The previous studies found longer TLs in Hispanic adults compared to white adults but no significant difference in infant TL.^32,33^ The longer TL in placentas associated with Latino mothers is the first time reported here, to our knowledge. Additionally, we also observe for the first time the association between maternal blood type O and shorter TL. In a systematic analysis of 9 studies, women with blood type O were found to have slightly less risk of preeclampsia than non-O blood type^34^. However, in our data blood type didn’t show significant interaction with severe preeclampsia. The connection between blood type and TL is still significant after controlling for preeclampsia. Future works are needed to explore more mechanisms of longer TL in Latinos and shorter TL associated with maternal blood type O. The potential missing components are the baby’s blood type and ethnicities which are not reported here.

### Strength and limitations

The study addressed the possible (or lack of) associations between placental TL with PE and other physiological and clinical phenotypes, using the largest multiracial cohort so far. In particular, the subjects include relatively large Asian (n = 133) and Pacific Islander populations (n=59), who were less studied in previous research compared to other ethnicities. One limitation is that the compositions of the cell types and their telomere length differences are also other possible sources of variations. As the placenta biology is stepping into single cell era,^7,35^ technologies such as single cell sequencing that identifies cell types and proportions, and ΩqPCR method that determines absolute telomere length from single-cell level,^36^ could reveal the heterogeneity of TL that are masked by the current tissue level measurements.

## Conclusions

In conclusion, our study showed no significant telomere change in PE complicated placentas compared to healthy controls. PE, though linked to stress and alteration in placentas, does not appear to lead to significant damage in placental telomere length, a biomarker for placenta aging.

## Supporting information

All supplementary tables and figures

## Data Availability

All data produced in the present study are available upon reasonable request to the authors

## ACKNOWLEDGEMENT

We thank Jason Delos Reyes from the Bioinformatics Core for matching the cases and control samples from the biobank. This research was supported by grants K01ES025434 awarded by NIEHS through funds provided by the trans-NIH Big Data to Knowledge (BD2K) initiative (www.bd2k.nih.gov), P20 COBRE GM103457 awarded by NIH/NIGMS, R01 LM012373 and R01 LM012907 awarded by NLM, and R01 HD084633 awarded by NICHD to L.X. Garmire.

