## Supplementary material for "Placental telomere length shortening is not associated with preeclampsia but the gestational age": All supplementary tables and figures

**Supplementary Materials**

**Supplementary Table 1: Full model linear regression results using 20 variables and 249 samples**

|  | ***Beta value*** | ***Standard Error*** | ***P-value*** | ***Significance*** |
| --- | --- | --- | --- | --- |
| ***(Intercept)*** | *6.86* | *0.77* | *2.52E-16* | ***** |
| ***Mothers; Age*** | *0.0054* | *0.0044* | *0.22* |  |
| ***Gestational Age*** | *-0.039* | *0.017* | *0.019* | *** |
| ***Membrane Ruptured*** | *-0.019* | *0.059* | *0.74* |  |
| ***Vaginal Bleeding*** | *0.095* | *0.071* | *0.18* |  |
| ***Weight Gain*** | *0.00029* | *0.0016* | *0.86* |  |
| ***Mothers' Height*** | *-0.017* | *0.066* | *0.80* |  |
| ***BMI*** | *-0.0016* | *0.0043* | *0.71* |  |
| ***Chronic Hypertension*** | *-0.0081* | *0.078* | *0.92* |  |
| ***History of Asthma*** | *-0.090* | *0.070* | *0.20* |  |
| ***History of Anemia*** | *-0.011* | *0.070* | *0.88* |  |
| ***Baby sex male*** | *-1.36* | *0.67* | *0.045* | *** |
| ***Sample Group Controls*** | *-0.095* | *0.074* | *0.20* |  |
| ***Blood type O vs non-O*** | *-0.053* | *0.10* | *0.60* |  |
| ***Blood type A vs non-A*** | *0.075* | *0.10* | *0.47* |  |
| ***Blood type B vs non-B*** | *0.028* | *0.11* | *0.79* |  |
| ***Asian vs non-Asian*** | *0.12* | *0.21* | *0.56* |  |
| ***Caucasian vs non-Caucasian*** | *0.054* | *0.22* | *0.80* |  |
| ***Latin vs non-Latin*** | *0.35* | *0.22* | *0.12* |  |
| ***Pacific Islander vs non-Pacific Islander*** | *0.18* | *0.21* | *0.38* |  |
| ***Macrosomia*** | *-0.14* | *0.12* | *0.23* |  |
| ***Gestational Age:Baby sex male*** | *0.036* | *0.018* | *0.049* | *** |

**Supplementary Table 2: Linear regression result on full-term (gestational age > 38 week) samples(n=158)**

| ***Variable Name*** | ***Beta*** | ***Standard Error*** | ***P-value*** | ***Significance*** |
| --- | --- | --- | --- | --- |
| ***(Intercept)*** | *7.68E+00* | *1.65E+00* | *7.36E-06* | ***** |
| ***Mothers' Age*** | *1.41E-03* | *5.44E-03* | *0.80* |  |
| ***Gestational Age*** | *-4.92E-02* | *3.96E-02* | *0.22* |  |
| ***Membrane Ruptured*** | *1.96E-03* | *6.67E-02* | *0.98* |  |
| ***Vaginal Bleeding*** | *8.55E-02* | *7.60E-02* | *0.26* |  |
| ***Weight Gain*** | *-1.88E-05* | *1.97E-03* | *0.99* |  |
| ***Mothers' Height*** | *-6.43E-02* | *9.21E-02* | *0.49* |  |
| ***BMI*** | *-3.84E-03* | *5.90E-03* | *0.52* |  |
| ***Chronic Hypertension*** | *8.42E-02* | *1.26E-01* | *0.50* |  |
| ***History of Asthma*** | *-1.86E-01* | *8.91E-02* | *0.039* | *** |
| ***History of Anemia*** | *1.58E-01* | *9.59E-02* | *0.10* |  |
| ***Baby sex male*** | *3.94E-02* | *6.59E-02* | *0.55* |  |
| ***Sample Group: preeclampsia*** | *-4.34E-02* | *9.42E-02* | *0.65* |  |
| ***Blood type O*** | *-1.32E-01* | *1.21E-01* | *0.28* |  |
| ***Blood type A*** | *-1.27E-02* | *1.24E-01* | *0.92* |  |
| ***Blood type B*** | *-4.09E-02* | *1.35E-01* | *0.76* |  |
| ***Asian vs non-Asian*** | *3.55E-02* | *2.43E-01* | *0.88* |  |
| ***Caucasian vs non-Caucasian*** | *1.26E-01* | *2.49E-01* | *0.61* |  |
| ***Latin vs non-Latin*** | *1.24E-01* | *2.56E-01* | *0.63* |  |
| ***Pacific Islander vs non-Pacific Islander*** | *7.49E-02* | *2.43E-01* | *0.76* |  |
| ***Macrosomia*** | *-1.03E-01* | *1.23E-01* | *0.41* |  |

**Supplementary Figure 1: The Pearson’s correlation heatmap of all 20 variables.**

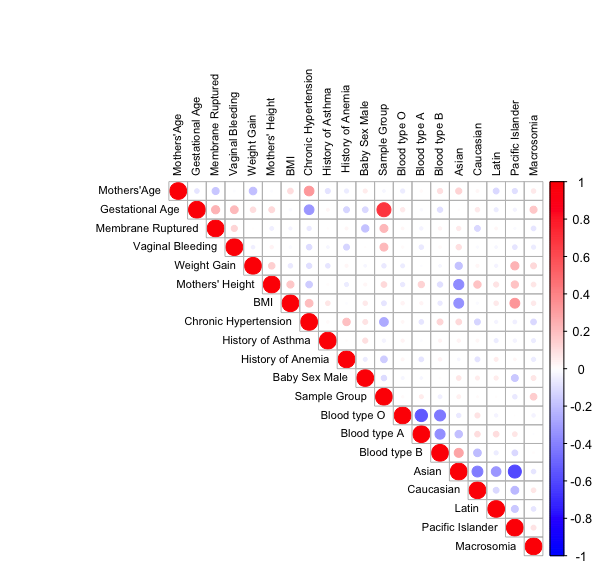

**Supplementary Figure 2: The histograms and density plots of placental TL before and after log-transformation.**

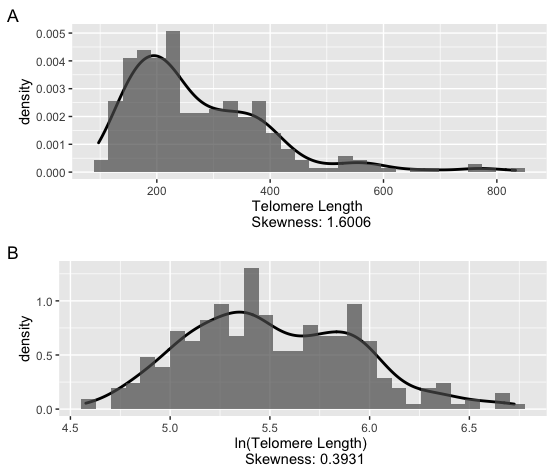
